# ICU Outcomes and Survival in Patients with Severe COVID-19 in the Largest Health Care System in Central Florida

**DOI:** 10.1101/2020.08.25.20181909

**Authors:** Eduardo Oliveira, Amay Parikh, Arnaldo Lopez-Ruiz, Maria Carrilo, Joshua Goldberg, Martin Cearras, Khaled Fernainy, Sonja Andersen, Luis Mercado, Jian Guan, Hammad Zafar, Patricia Louzon, Amy Carr, Natasha Baloch, Richard Pratley, Scott Silverstry, Vincent Hsu, Jason Sniffen, Victor Herrera, Neil Finkler

## Abstract

**Background:** Observational studies have consistently described poor clinical outcomes and increased ICU mortality in patients with severe coronavirus disease 2019 (COVID-19) who require mechanical ventilation (MV). Our study describes the clinical characteristics and outcomes of patients with severe COVID-19 admitted to ICU in the largest health care system in the state of Florida, United States.

**Methods:** Retrospective cohort study of patients admitted to ICU due to severe COVID-19 in AdventHealth health system in Orlando, Florida from March 11^th^ until May 18th, 2020. Patients were characterized based on demographics, baseline comorbidities, severity of illness, medical management including experimental therapies, laboratory markers and ventilator parameters. Major clinical outcomes analyzed at the end of the study period were: hospital and ICU length of stay, MV-related mortality and overall hospital mortality of ICU patients.

**Results:** Out of total of 1283 patients with COVID-19, 131 (10.2%) met criteria for ICU admission (median age: 61 years [interquartile range {IQR}, 49.5-71.5]; 35.1% female). Common comorbidities were hypertension (84; 64.1%), and diabetes (54; 41.2%). Of the 131 ICU patients, 109 (83.2%) required MV and 9 (6.9%) received ECMO. Lower positive end expiratory pressure (PEEP) were observed in survivors [9.2 (7.7-10.4)] vs non-survivors [10 (9.1-12.9] p= 0.004]. Compared to non-survivors, survivors had a longer MV length of stay (LOS) [14 (IQR 8-22) vs 8.5 (IQR 5-10.8) p< 0.001], Hospital LOS [21 (IQR 13-31) vs 10 (7-1) p< 0.001] and ICU LOS [14 (IQR 7-24) vs 9.5 (IQR 6-11), p < 0.001]. The overall hospital mortality and MV-related mortality were 19.8% and 23.8% respectively. After exclusion of hospitalized patients, the hospital and MV-related mortality rates were 21.6% and 26.5% respectively.

**Conclusions:** Our study demonstrates an important improvement in mortality of patients with severe COVID-19 who required ICU admission and MV in comparison to previous observational reports and emphasize the importance of standard of care measures in the management of COVID-19.

## Introduction

Coronavirus disease 2019 (COVID-19) have affected over 7 million of people around the world since December 2019 [1] and in the United States has resulted so far in more than 100,000 deaths [2]. Epidemiological studies have shown that 6 to 10% of patients develop a more severe form of COVID-19 and will require admission to the intensive care unit (ICU) due to acute hypoxemic respiratory failure [3]. Most of these patients admitted to ICU, will finally require invasive mechanical ventilation (MV) due to diffuse lung injury and acute respiratory distress syndrome (ARDS). Until now, most of the ICU reports from United States have shown that severe COVID-19-associated ARDS (CARDS) is associated with prolonged MV and increased mortality [4]. In fact, retrospective and prospective case series from China and Italy have provided insight about the clinical course of severely ill patients with CARDS in which it demonstrates that extrapulmonary complications are also a strong contributor for poor outcomes [5,6]. In United States, population dense areas such as New York City, Seattle and Los Angeles have had the highest rates of infection resulting in significant overload to hospitals and ICU systems [2,7,8]. However, tourist destinations and areas with a large elderly population like the state of Florida pose a remaining concern for increasing infection rates that may lead to high national mortality.

Mortality rates reported in patients with severe COVID-19 in the ICU range from 50-65% [7, 8, 9]. In patients requiring MV, mortality rates have been reported to be as high as 97% [10]. Regional experiences in the management of critically ill patients with severe COVID-19 have varied between cities and countries, and recent reports suggest a lower mortality rate [11]. The regional and institutional variations in ICU outcomes and overall mortality are not clearly understood yet and are not related to the use experimental therapies, given the fact that recent reports with the use remdesivir [12], hydroxychloroquine/azithromycin [13], lopinavir-ritonavir [14] and convalescent plasma [15,16] have been inconsistent in terms of mortality reduction and improvement of ICU outcomes. Therefore, the poor ICU outcomes and high mortality rate observed during CARDS have raised concerns about the strategies of mechanical ventilation and the success in delivering standard of care measures.

Our observational study is so far the first and largest in the state of Florida to describe the demographics, baseline characteristics, medical management and clinical outcomes observed in patients with CARDS admitted to ICU in a multihospital health care system.

## Patient and Methods

Retrospective cohort study conducted at AdventHealth Central Florida Division (AHCFD), the largest health system in central Florida. AHCFD is comprised of 9 hospitals with a total of 2885 beds servicing the 8 million residents of Orange County and surrounding regions. All patients with COVID-19 who met criteria for critical care admission from AdventHealth hospitals were transferred and managed at AdventHealth Orlando, a 1368-bed hospital with 170 ICU beds and dedicated inhouse 24/7 intensivist coverage. This study was approved by the institutional review board of AHCFD. All critical care admissions from March 11 to May 18, 2020 presenting to any one of the 9 AHCFD hospitals were included. All consecutive critically ill patients had confirmed severe acute respiratory syndrome coronavirus 2 (SARS-CoV-2) infection by positive result on polymerase chain reaction (PCR) testing of a nasopharyngeal sample or tracheal aspirate. Due to some of the documented shortcomings of PCR testing early in this pandemic, some patients required more than one test to document positivity. Clinical outcomes of the included population were monitored until May 27, 2020, the final date of study follow-up. All critically ill COVID-19 patients were assigned in 2 ICUs with a total capacity of 80 beds. Patients not requiring ICU level care were admitted to a specially dedicated isolation unit at each AHCFD hospital. Standardized respiratory care was implemented favoring intubation and MV over non-invasive positive pressure ventilation. The ICUs employed dedicated respiratory therapists, with extensive training in the care of patients with ARDS.

We considered the following criteria to admit patients to ICU: 1) Oxygen saturation (O2 sat) less than 93% on more than 6 liters oxygen (O2) via nasal cannula (NC) or PO2 < 65 mmHg with 6 liters or more O2, or respiratory rate (RR) more than 22 per minute on 6 liters O2, 2) PO2/FIO2 ratio less than 300, 3) any patient with positive PCR test for SARS-CoV-2 already on requiring MV or with previous criteria. We accomplished strict protocol adherence for low tidal volume ventilation targeting a plateau pressure goal of less than 30 cmH2O and a driving pressure of less than 15 cmH2O. We followed ARDS network low PEEP, high FiO2 table in the majority of our cases [17]. Those patients requiring mechanical ventilation were supervised by board-certified critical care physicians (intensivists). Intensivist were not responsible for more than 20 patients per 12 hours shift. Nursing did not exceed ratios of one nurse to two patients. Early paralysis and prone positioning were achieved with the assistance of a dedicated prone team. Prone Positioning techniques were consistent with the PROSEVA trial recommendations [18]. The 30 ml/kg crystalloid resuscitation recommendation was applied for those patients presenting with evidence of septic shock and fluid resuscitation was closely monitored to minimize overhydration [19]. Based on recent reports showing hypercoagulable state and increased risk of thrombosis in patients with COVID-19, deep vein thrombosis (DVT) prophylaxis was initiated by following an institutional algorithm that employed D-dimer levels and rotational thromboelastometry (ROTEM) to determine the risk of thrombosis [20]. Prophylactic anticoagulation ranged from unfractionated heparin at 5000 units subcutaneously (SC) every eight hours or enoxaparin 0.5 mg/kg SC daily to full anticoagulation with either an unfractionated heparin infusion or enoxaparin 1 mg/kg SC twice daily.

A selected number of patients received remdesivir as part of the expanded access or compassionate use programs, as well as through the Emergency Use Authorization (EUA) supply distributed by the Florida Department of Health. Patients were also enrolled in institutional review board (IRB) approved studies for convalescent plasma and other COVID-19 investigational treatments.

Data were collected from the enterprise electronic health record (Cerner; Cerner Corp. Kansas City, MO) reporting database, and all analyses were performed using version 3.6.3 of the R programming language (R Project for Statistical Computing; R Foundation). Patients were considered to have confirmed infection if the initial or repeat test results were positive. Repeat tests were performed after an initial negative test by obtaining a lower respiratory sample if there was a high clinical pretest probability of COVID-19. Transfers between system hospitals were considered a single visit.

Data collected included patient demographic information, comorbidities, triage vitals, initial laboratory tests, inpatient medications, treatments (including invasive mechanical ventilation and renal replacement therapy), and outcomes (including length of stay, discharge, readmission, and mortality). All clinical outcomes are presented for patients who were admitted to the cohort ICU during the study period (discharged alive, remained in the hospital or dead). Clinical outcomes available at the study end point are presented, including invasive mechanical ventilation, ICU care, renal replacement therapy, and hospital length of stay. Race data were self-reported within prespecified fixed categories. Initial laboratory testing was defined as the first test results available, typically within 24 hours of admission. For initial laboratory testing and clinical studies for which not all patients had values, percentages of total patients with completed tests are shown. The scores APACHE IVB, MEWS, and SOFA scores were computed to determine the severity of illness and data for these scoring was provided by the electronic health records. The. predicted hospital mortalities were calculated using the equations of APACHE IVB utilizing principal diagnosis of viral and bacterial pneumonia [21].

Patient characteristics and clinical outcomes were compared based on survival status of COVID-19 positive patients. Categorical fields are displayed as percentages and continuous fields are presented as means or standard deviations (SD) or median and interquartile range. Bivariate analysis was performed by survival status of COVID-19 positive patients to examine differences in the survival and non-survival group using chi-square tests and Welch’s t-test.

## Results

During March 11 to May 18, a total of 1283 COVID-19 positive patients were evaluated in the Emergency Department or ambulatory care centers of AHCFD. Out of 1283, 429 (33.4%) were admitted to AHCFD hospitals, of which 131 (30.5%) were admitted to the AdventHealth Orlando COVID-19 ICU. This result suggests a 10.2% (131/1283) rate of ICU admission (Figure 1 - Study Diagram).

**Figure 1.**
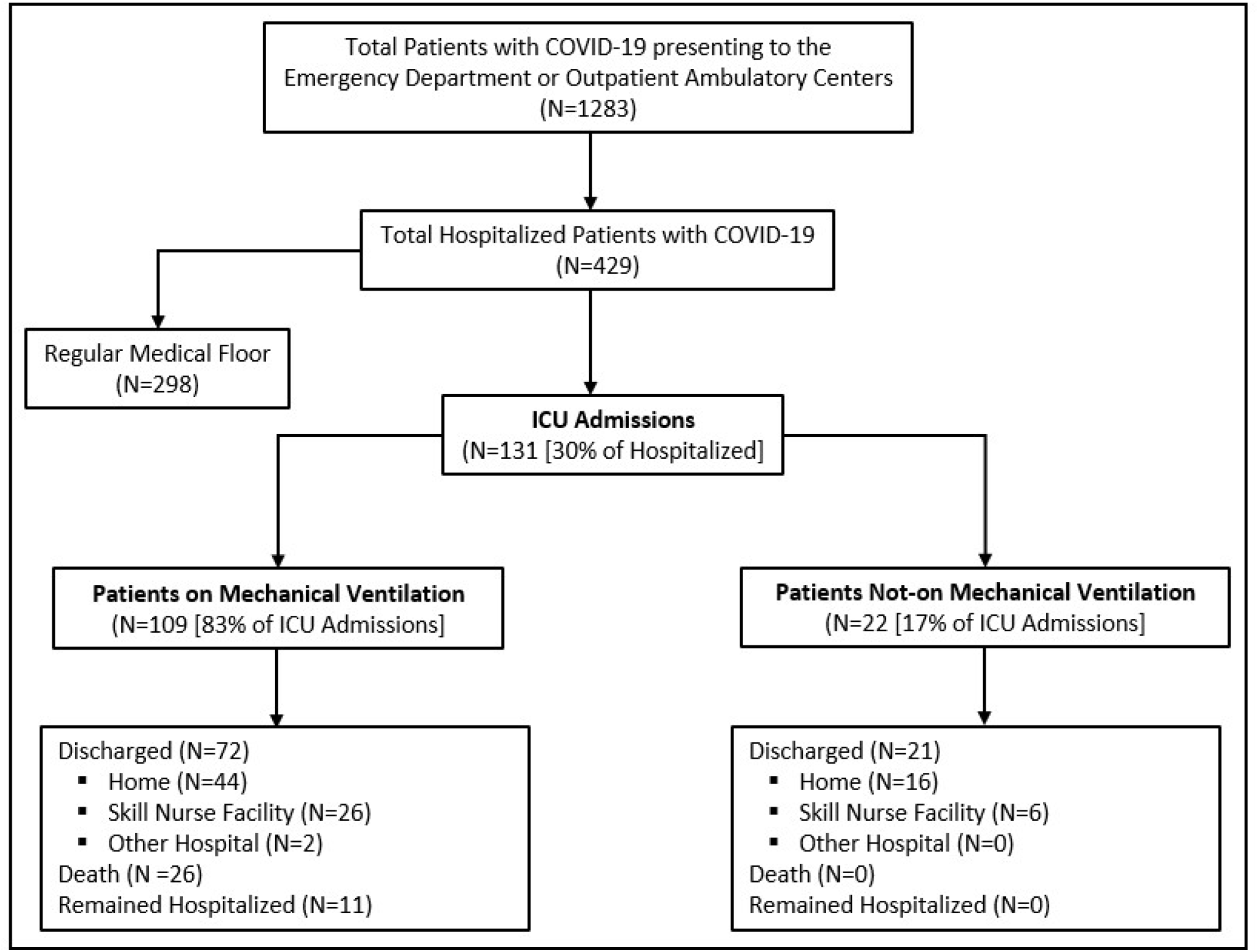
Study Flow Diagram of Patients with COVID-19 admitted to Intensive Care Unit (ICU)

Baseline demographic and clinical characteristics of patients are summarized in Table 1 and Table 2 respectively. The median age of the patients admitted to the ICU was 61 (IQR 49.5-71.5). Deceased patients were older with a median age of 71.5 (IQR 62-80, p <0.001). A majority of patients were male (64.9%), 15 (11%) were black, and the majority of patients were classified as white and other (116, 88.5%). Hypertension was the most common co-morbid condition (84 pts, 64%), followed by diabetes (54, 41%) and coronary artery disease (21, 16%). Evidence of heart failure, chronic kidney disease (CKD) and dementia were associated with non-survivors. Obesity (BMI 30-39.9) was observed in 50 patients (38.2%), and 7 (5.3%) patients had a BMI of 40 or greater. Approximately half of the study population had commercial insurance (67, 51%) followed by Medicare (40, 30.5%), Medicaid (12, 9.2%) and uninsured (12, 9.2%). Initial presentation with Oxygen (O2) saturation < 90% (p= 0.006), respiratory rate > 22 (p= 0.003) and systolic blood pressure < 90mmhg (p= 0.008) were more commonly present in non-survivors.

**Table 1.** Baseline Demographic Characteristics of the Patients Admitted to ICU with COVID-19.

| Patient Characteristics<br>[n (%) or Median (IQR)] | Total Patients<br>(n=131) | Patients Alive<br>(n=105) | Patients<br>Deceased (n=26) | *p-value < 0.05 |
| --- | --- | --- | --- | --- |
| **Age (years)** | 61 [49.5-71.5] | 60 [49-69] | 71.5 [62-80] | *0.001 |
| 18 o <55 | 41 (31.3 %) | 38 (36.2 %) | 3 (11.5 %) | *0.003 |
| 55-64 | 36 (27.5 %) | 31 (29.5 %) | 5 (19.2 %) | 0.262 |
| 65-74 | 34 (26.0 %) | 26 (24.8 %) | 8 (30.8 %) | 0.558 |
| ≥ 75 | 20 (15.3 %) | 10 (9.5 %) | 10 (38.5 %) | *0.008 |
| **Gender** |  |  |  |  |
| Male | 46 (35.1%) | 36 (34.2 %) | 10 (38.5 %) | 0.701 |
| Female | 85 (64.9 %) | 69 (65.7 %) | 16 (61.5%) | 0.701 |
| **Race** |  |  |  |  |
| African American | 15 (11.5 %) | 12 (11.4 %) | 3 (11.5%) | 0.988 |
| Caucasian | 59 (45 %) | 45 (42.9 %) | 14 (53.8 %) | 0.328 |
| Other | 57 (43.5 %) | 48 (45.7 %) | 9 (34.6 %) | 0.306 |
| **Medical Insurance** |  |  |  |  |
| Commercial | 67 (51.1 %) | 58 (55.2 %) | 9 (34.6 %) | 0.061 |
| Medicare | 40 (30.5 %) | 26 (24.8 %) | 14 (53.8 %) | *0.011 |
| Medicaid | 12 (9.2 %) | 10 (9.5 %) | 2 (7.7 %) | 0.764 |
| Uninsured | 12 (9.2 %) | 11 (10.5 %) | 1 (3.9 %) | 0.179 |
| **Body Mass Index (BMI)** |  |  |  |  |
| ≤ 25 | 40 (30 %) | 33 (31.4 %) | 7 (26.9 %) | 0.654 |
| ≥ 40 | 7 (5.3 %) | 5 (4.8 %) | 2 (7.7 %) | 0.612 |
| **Comorbidities** |  |  |  |  |
| Diabetes | 54 (41.2 %) | 39 (37.1 %) | 15 (57.7 %) | 0.092 |
| Hypertension | 84 (64.1 %) | 65 (61.9 %) | 19 (73.1 %) | 0.404 |
| Cerebrovascular disease | 7 (5.3 %) | 4 (3.8 %) | 3 (11.5 %) | 0.104 |
| Heart Failure | 12 (9.2 %) | 4 (3.8 %) | 8 (30.8 %) | *0.001 |
| CKD stage ≥ 3 | 14 (10.7 %) | 5 (4.8 %) | 9 (34.6 %) | *0.001 |
| Dementia | 10 (7.6 %) | 5 (4.8 %) | 5 (19.2 %) | *0.026 |
| Malignancy | 9 (6.9 %) | 9 (8.6 %) |  | 0.203 |
| **Smoking History** |  |  |  |  |
| Current/Former smoker | 23 (17.6 %) | 16 (15.2 %) | 7 (26.9 %) | 0.119 |
| Non-smoker | 35 (26.7 %) | 83 (80.6 %) | 17 (65.4 %) | 0.229 |

**Table 2.** Baseline Clinical Characteristics of the Patients Admitted to ICU with COVID-19.

| Clinical Characteristics<br>[n (%) or Median (IQR)] | Total Patients<br>(n=131) | Patients Alive<br>(n=105) | Patients<br>Deceased (n=26) | *p-value < 0.05 |
| --- | --- | --- | --- | --- |
| **Initial Vital Signs** |  |  |  |  |
| Temperature > 38 °C | 53 (40.5 %) | 44 (41.9 %) | 9 (34.6 %) | 0.649 |
| Oxygen Saturation < 90% | 35 (26.7 %) | 22 (21.0 %) | 13 (50 %) | *0.006 |
| Supplemental Oxygen at Admission Triage | 65 (49.6 %) | 49 (46.7 %) | 16 (61.5 %) | 0.255 |
| Respiratory Rate > 22/min | 54 (41.2 %) | 36 (34.3 %) | 18 (69.2 %) | *0.003 |
| Heart Rate > 100/min | 56 (42.7 %) | 48 (45.7 %) | 8 (30.8 %) | 0.247 |
| Systolic Blood Pressure (SBP) ≤ 90 mmHg | 11 (8.4 %) | 5 (4.76 %) | 6 (23.1 %) | *0.008 |
| **Onset of symptoms-to-Hospital Admission (Days)** | 5.5 [3.0-8.3] | 7 [4-10] | 3.5 [2-6.3] | *0.03 |
| **Severity Score** |  |  |  |  |
| MEWS score | 8 [5-10] | 7 [5-10] | 10 [7.3-11] | *0.003 |
| SOFA score | 3 [2-5] | 3 [2-4] | 5.5 [3.3-7.8] | *0.001 |
| APACHE IVB score | 50.5 [37-66] | 47 [34.8-61] | 69 [59.3-77.5] | *0.001 |
| **Initial Laboratory Results** |  |  |  |  |
| Absolute Neutrophil Count (x10<sup>3</sup>/UL) | 0.85 [0.58-1.10] | 0.84 [0.57-1.09] | 0.89 [0.66-1.16] | 0.624 |
| ALT (Units/L) | 30 [20-50.5] | 27 [19-42] | 37 [30-75.8] | 0.630 |
| AST (Units/L) | 44 [30-61] | 38 [29-60] | 54 [45-74.8] | 0.865 |
| Bilirubin Total (mg/dl) | 0.4 [0.3-0.7] | 0.4 [0.3-0.7] | 0.4 [0.3-0.6] | 0.172 |
| Creatinine (mg/dl) | 1.0 [0.8-1.3] | 1.1 [0.9-1.4] | 0.9 [0.7-1.0] | 0.615 |
| C-Reactive Protein (mg/L) | 115 [59-186.3] | 116 [62.4-186.2] | 105 [41.3-188.2] | 0.784 |
| D-Dimer (µg/DL) | 1.4 [0.8-3.2] | 1.4 [0.8-3.3] | 1.7 [0.8-3.2] | 0.948 |
| Ferritin (ng/ml) | 848 [441-1541] | 910 [488-1557] | 544 [333-1494] | 0.855 |
| Interleukin-6 [IL-6] (pg/ml) | 18 [7.0-46.5] | 21 [9-48] | 11.5 [5-33.8] | 0.281 |
| Lactic acid (mmol/L) | 1.4 [1.1-1.7] | 1.4 [1.1-1.7] | 1.5 [1.0-1.9] | 0.652 |
| Procalcitonin (ng/ml) | 0.25 [0.15-0.67] | 0.25 [0.15-0.68] | 0.26 [0.15-0.61] | 0.701 |
| Prothrombin Time (seconds) | 14 [13.0-15.1] | 14 [13.1-14.9] | 14 [12.9-15.2] | 0.649 |
| Troponin-T (ng/ml) | 0.01 [0.01-0.02] | 0.01 [0.01-0.02] | 0.01 [0.01-0.02] | 0.329 |

Risk adjusted severity (SOFA, MEWS, APACHE IVB) scores were significantly higher in non-survivors (p< 0.003). Median C-reactive protein on hospital admission was 115 mg/L (IQR 59.3-186.3; upper limit of normal 5 mg/L), median Ferritin was 848 ng/ml (IQR 441-1541); upper limit of normal 336 ng/ml), D-dimer was 1.4 ug/mL (IQR 0.8-3.2; upper limit of normal 0.8 ug/mL), and IL-6 level was 18 pg/mL (IQR 7-46.5; upper limit of normal 2 pg/mL). No significant differences in the laboratory and inflammatory markers were observed between survivors and non-survivors.

ICU specific management and interventions including experimental therapies and hospital as well as ICU length of stay (LOS) are described in table 3. There were 109 patients (83%) who received MV. Compared to non-survivors, survivors had a longer time on the ventilator [14 (IQR 8-22) versus 8.5(IQR 5-10.8) p< 0.001], Hospital LOS [21 (IQR 13-31) versus 10 (7-1) p< 0.001] and ICU LOS [14 (IQR 7-24) versus 9.5 (IQR 6-11), p < 0.001]. Prone positioning was performed in 46.8% of the study subjects and 77% of the mechanically ventilated patients received neuromuscular blockade to improve hypoxemia and ventilator synchrony. Average PaO2/FiO2 during hospitalization was lower in non-survivors [167 (IQR 132.7-194.1)] versus survivors [202 (IQR 181.8-234.4)] p< 0.001. Lower positive end expiratory pressure (PEEP) averages were observed in survivors [9.2 (7.7-10.4)] vs non-survivors [10 (9.1-12.9] p= 0.004]. Median Driving pressure were similar between the two groups (12.7 [10.8-15.1)].

**Table 3.**
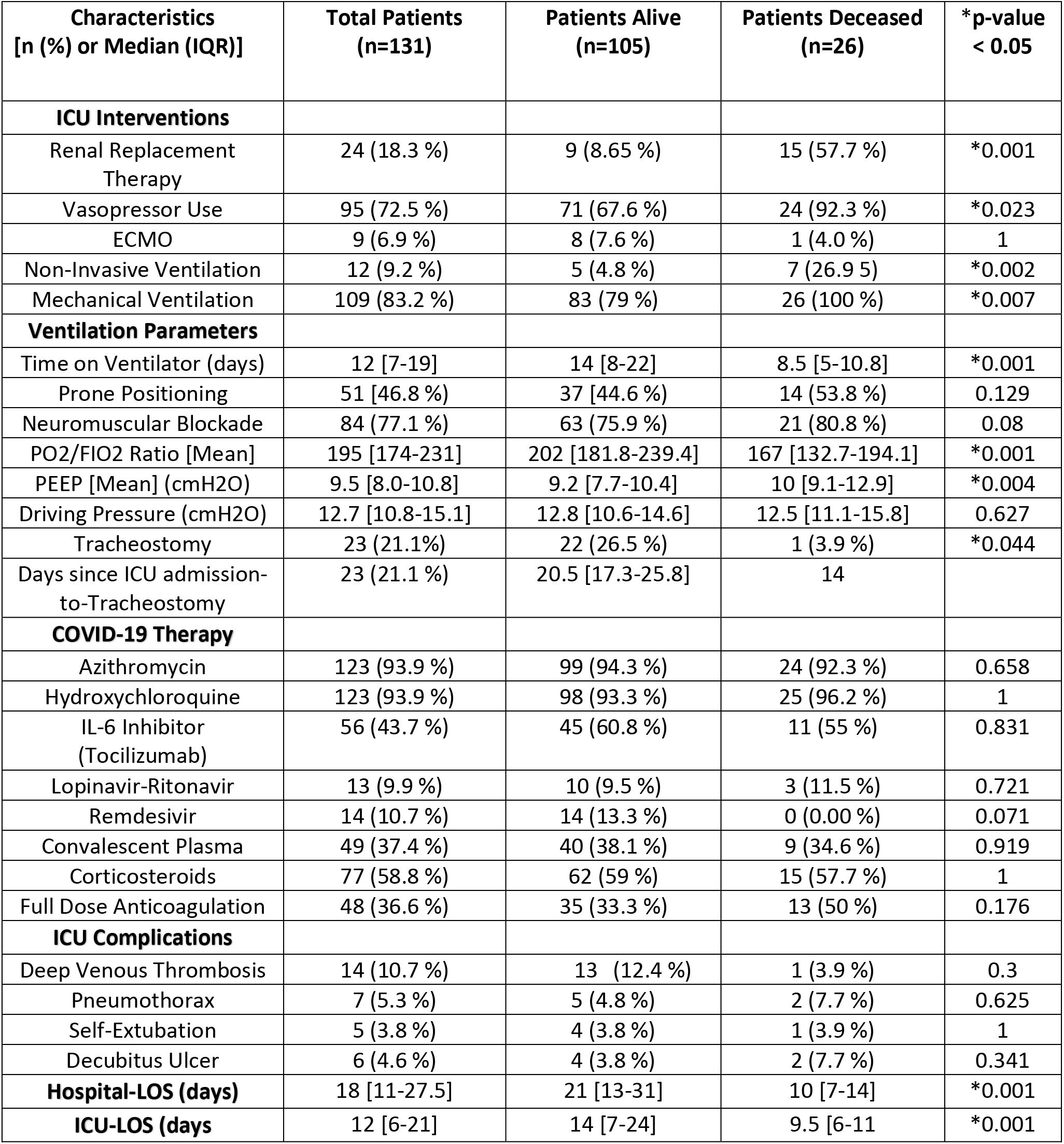
ICU Management, Interventions and Length of Stay (LOS) of Patients with COVID-19.

| **Characteristics**<br>**[n (%) or Median (IQR)]** | **Total Patients**<br>**(n=131)** | **Patients Alive**<br>**(n=105)** | **Patients Deceased**<br>**(n=26)** | **\*p-value**<br>**< 0.05** |
| --- | --- | --- | --- | --- |
| **ICU Interventions** |  |  |  |  |
| Renal Replacement Therapy | 24 (18.3 %) | 9 (8.65 %) | 15 (57.7 %) | *0.001 |
| Vasopressor Use | 95 (72.5 %) | 71 (67.6 %) | 24 (92.3 %) | *0.023 |
| ECMO | 9 (6.9 %) | 8 (7.6 %) | 1 (4.0 %) | 1 |
| Non-Invasive Ventilation | 12 (9.2 %) | 5 (4.8 %) | 7 (26.9 %) | *0.002 |
| Mechanical Ventilation | 109 (83.2 %) | 83 (79 %) | 26 (100 %) | *0.007 |
| **Ventilation Parameters** |  |  |  |  |
| Time on Ventilator (days) | 12 [7-19] | 14 [8-22] | 8.5 [5-10.8] | *0.001 |
| Prone Positioning | 51 [46.8 %] | 37 [44.6 %] | 14 (53.8 %) | 0.129 |
| Neuromuscular Blockade | 84 (77.1 %) | 63 (75.9 %) | 21 (80.8 %) | 0.08 |
| PO2/FIO2 Ratio [Mean] | 195 [174-231] | 202 [181.8-239.4] | 167 [132.7-194.1] | *0.001 |
| PEEP [Mean] (cmH2O) | 9.5 [8.0-10.8] | 9.2 [7.7-10.4] | 10 [9.1-12.9] | *0.004 |
| Driving Pressure (cmH2O) | 12.7 [10.8-15.1] | 12.8 [10.6-14.6] | 12.5 [11.1-15.8] | 0.627 |
| Tracheostomy | 23 (21.1%) | 22 (26.5 %) | 1 (3.9 %) | *0.044 |
| Days since ICU admission-to-Tracheostomy | 23 (21.1 %) | 20.5 [17.3-25.8] | 14 |  |
| **COVID-19 Therapy** |  |  |  |  |
| Azithromycin | 123 (93.9 %) | 99 (94.3 %) | 24 (92.3 %) | 0.658 |
| Hydroxychloroquine | 123 (93.9 %) | 98 (93.3 %) | 25 (96.2 %) | 1 |
| IL-6 Inhibitor (Tocilizumab) | 56 (43.7 %) | 45 (60.8 %) | 11 (55 %) | 0.831 |
| Lopinavir-Ritonavir | 13 (9.9 %) | 10 (9.5 %) | 3 (11.5 %) | 0.721 |
| Remdesivir | 14 (10.7 %) | 14 (13.3 %) | 0 (0.00 %) | 0.071 |
| Convalescent Plasma | 49 (37.4 %) | 40 (38.1 %) | 9 (34.6 %) | 0.919 |
| Corticosteroids | 77 (58.8 %) | 62 (59 %) | 15 (57.7 %) | 1 |
| Full Dose Anticoagulation | 48 (36.6 %) | 35 (33.3 %) | 13 (50 %) | 0.176 |
| **ICU Complications** |  |  |  |  |
| Deep Venous Thrombosis | 14 (10.7 %) | 13 (12.4 %) | 1 (3.9 %) | 0.3 |
| Pneumothorax | 7 (5.3 %) | 5 (4.8 %) | 2 (7.7 %) | 0.625 |
| Self-Extubation | 5 (3.8 %) | 4 (3.8 %) | 1 (3.9 %) | 1 |
| Decubitus Ulcer | 6 (4.6 %) | 4 (3.8 %) | 2 (7.7 %) | 0.341 |
| **Hospital-LOS (days)** | 18 [11-27.5] | 21 [13-31] | 10 [7-14] | *0.001 |
| **ICU-LOS (days)** | 12 [6-21] | 14 [7-24] | 9.5 [6-11] | *0.001 |

Vasopressors were required in 72.5% of the ICU patients (non-survivors 92.3% versus survivors 67.6%, p= 0.023). Renal replacement therapy was required in 24 (18%), out of which 15 patients (57.7%) expired. Only 9 of 131 ICU patients, received extracorporeal membrane oxygenation (ECMO), with most of them surviving (8, 88%). Of these 9 patients, 8 were treated with veno-venous ECMO (survival 7 of 8) and one with veno-arterial-venous ECMO (survival 1 of 1).

The majority of patients (N=123, 93.9%) received a combination of azithromycin and hydroxychloroquine. Tocilizumab was utilized in 56 (43.7%), and 37 (28.2%) were enrolled in blinded placebo-controlled studies aimed at the inflammatory cascade. Convalescent plasma was administered in 49 (37.4%) patients. A total of 14 (10.7%) received remdesivir via expanded access or compassionate use programs, as well as through the Emergency Use Authorization (EUA) supply distributed by the Florida Department of Health. Full anticoagulation was given to 48 (N=131, 36.6%) of the patients and 77 (N=131, 58.8%) received high dose corticosteroids (methylprednisolone 40mg every 8 hours for 7 days or dexamethasone 20 mg every day for 5 days followed by 10 mg every day for 5 days). Of the 109 patients requiring mechanical ventilation, 61 (55%) received the previously mentioned dose of methylprednisolone or dexamethasone. Although our study was not designed to assess the effectiveness of any of the above medications, no significant differences between survivors and non-survivors were observed through bivariate analysis.

ICU outcomes at the end of study period are described in Table 4. Of the total amount of patients admitted to ICU (N=131), 80.2% (N=105) remained alive at the end of the study period. Of those alive patients, 89.5% (N=94) were discharged from the hospital. Of these patients who were discharged, 60 (45.8%) went home, 32 (24.4%) were discharged to skill nurse facilities and 2 (1.5%) were discharged to other hospitals. During the study period, 26 patients of the total (N=131) expired (19.8% overall mortality). Excluding those patients who remained hospitalized (N=11 [8.4% of 131] at the end of study period, adjusted hospital mortality of ICU patients was 21.6%. Higher survival rate was observed in patients younger than 55 years old (p= 0.003) with the highest mortality rate observed in those patients older than 75 years (p= 0.008). Of the total ICU patients who required invasive mechanical ventilation (N=109 [83.2%]), 26 patients (23.8%) expired during the study period. When the mechanical ventilation-related mortality was calculated excluding those patients who remained hospitalized, this rate increased to 26.5%. The APACHE IVB score-predicted hospital and ventilator mortality was 17% and 21% respectively for patients with a discharge disposition. Due to lack of risk-adjusted APACHE predictions specifically for patients with COVID 19-induced acute respiratory failure, we considered acute respiratory failure secondary to usual viral and/or bacterial pneumonia as the principal diagnoses to determine expected rates of mortality.

**Table 4.** ICU Outcomes in Patients with COVID-19 and Predicted Mortality *[%] based on APACHE IVB score.

| **Outcomes** | **N (Percentage %)** |
| --- | --- |
| **Patient Disposition** | 131 |
| ➤ Remained Hospitalized | 11 (8.4%) |
| ➤ Discharged Home | 60 (45.8 %) |
| ➤ Discharged to Skill Nurse Facility | 32 (24.4 %) |
| ➤ Discharged to another hospital | 2 (1.5 %) |
| ➤ Death | 26 (19.8 %) |
| **Overall Hospital Mortality of ICU patients** | 19.8 % |
| **Hospital Mortality of ICU patients – Excluding hospitalized patients** | 21.6 % *[17 %] |
| **Mechanical Ventilation-related mortality** | 23.8 % |
| **Mechanical Ventilation-related mortality – Excluding hospitalized patients.** | 26.5 % *[21 %] |

## Discussion

Among 429 admissions during the study period, 131 were admitted to our ICU (30.5%). Patients referred to our center from outside our system included patients to be evaluated for Extracorporeal Membrane Oxygenation (ECMO) and patients who experienced delays in hospital level of care due to travel on cruise lines. These patients universally required a higher level of care than our average patient admission and may explain our slightly higher ICU admission rate as compared to the literature (22-27.4%) [11,22]

Reports of ICU mortality due to COVID-19 around the world and in the Unites States, in particular, have ranged from 20-62% [7]. In mechanically ventilated patients, mortality has ranged from 50-97%. Observations from Wuhan have shown mortality rates of approximately 52% in COVID-19 patients with ARDS [23]. Cohorts in New York have shown a mortality rate in the mechanically ventilated population as high as 88.1% [4]. Based on these high mortality rates, there has been speculation that this disease process is different than typical ARDS, suggesting that standard ARDS mechanical ventilation strategies may not be as effective in reducing lung injury [24]. However, both our in-hospital and mechanical ventilation mortality rates were significantly lower than what has been reported in the literature (Table 4). In fact, our data suggests that COVID-19-induced ARDS requiring mechanical ventilation has a similar if not lower mortality than what has been previously observed in ARDS due to other infectious etiologies [25].

There are several potential explanations for our study findings. Our lower mortality could be partially explained by our lower average patient age or higher proportion of Non-African Americans as some studies have suggested a higher mortality in the African American population [26]. However, in countries where the majority population were non-black (China, Italy, and other countries in Europe), a high mortality rate was also observed. Our study population also had a higher rate of commercial insurance, which may suggest an improved baseline health status which has been associated with an overall lower all-cause mortality [27]. An additional factor to be considered is our geographical location: the warmer climate and higher humidity experienced in central Florida, have been associated with a lower mortality of the disease [28]. It is unclear whether these or other environmental factors could also be associated with a lower virulence for COVID-19 in our region.

Other relevant factors that in our opinion are likely to have influenced our outcomes were that our healthcare delivery system was never overwhelmed. We were allowed time to adapt our facility infrastructure, recruit and retain proper staffing, cohort all critical ill patients in one location to enhance staff expertise and minimize variation, secure proper personal protective equipment, develop proper processes of care, and follow an increasing number of medical Society best practice recommendations [29]

Overall, we strictly followed standard ARDS and respiratory failure management. Investigational treatments of uncertain efficacy were utilized when supported by available evidence at the time. (Table 3). The majority of our patients throughout March and April 2020 received hydroxychloroquine and azithromycin. Although the effectiveness and safety of this regimen has been recently questioned [13]. Our observed mortality does not suggest a detrimental effect of such treatment. Reported cardiotoxicity associated with this regimen was mitigated by frequent ECG monitoring and close monitoring of electrolytes. Potential benefit has been described for remdesivir in reducing the duration of hospital LOS, but it has not been shown to improve patient survival, especially in the critically ill population [12]. In addition, 43% of our patients received tocilizumab and 28.2% where enrolled in a blinded clinical trial of investigational drugs targeting the inflammatory cascade. The theoretical benefit of blocking cytokines, specially interleukin-6 [IL-6], which is one of main mediators of the cytokine release syndrome, has not been shown at this time to improve mortality or other outcomes [30]. Also, of note, 37.4% of our study population received convalescent plasma, and larger studies are underway to understand its role in the treatment of severe COVID-19 [15,31]. Another potential aspect that may have contributed to reduce our MV-related mortality and overall mortality is the use of steroids. We are reporting that 55% of the patients who required mechanical ventilation received methylprednisolone or dexamethasone. This specific population and the impact of steroids in respiratory parameters, ventilator-free days and survival need to be further evaluated. The dose and duration of steroids were based on the study by Villar J. et al, that showed an improvement in survival in patients with ARDS after using dexamethasone [34,35]. Interestingly, only 6.9% of our study population was referred for ECMO, however our ECMO mortality was much lower than previously reported in the literature (11% compared to 94%) [32,33].

This report has several limitations. This was an observational study conducted at a single health care system in a confined geographic area thus limiting the generalizability of our results. As with all observational studies, it is difficult to ascertain causality with ICU therapies as opposed to an association that existed due to the patients’ clinical conditions. Additionally, when examining multiple factors associated with survival, potential confounders may remain unidentified without a multivariate regression analysis. Finally, additional unmeasured factors might have played a significant role in survival. Despite these limitations, our experience and results challenges previously reported high mortality rates. In fact, our mortality rates for mechanically ventilated COVID-19 patients were similar to APACHE IVB predicted mortality, which was based on critically ill patients admitted with respiratory failure secondary to viral and/or bacterial pneumonia. Our study demonstrates the possibility of better outcomes for COVID-19 associated with critical illness, including COVID-19 patients requiring mechanical ventilation.

### Conclusion

Our study is the first and the largest in the state Florida and probably one of the most encouraging in the United States to show lower overall mortality and MV-related mortality in patients with severe COVID-19 admitted to ICU compared to other previous cases series. Our study does not support the previously reported overwhelmingly poor outcomes of mechanically ventilated patients with COVID-19 induced respiratory failure and ARDS. In fact, it is reassuring that the application of well-established ARDS and mechanical ventilation strategies can be associated with mortality and outcomes comparable to non-COVID-19 induced sepsis or ARDS.

## Data Availability

Availability of supporting data: Unidentified and analyzed numerical data of all the variables and clinical outcomes that are reported in this article can be provided to the editorial board and statistics specialist of the journal upon request.

## Acknowledgements

We would like to acknowledge the following AdventHealth Critical Care Consortium Research Group Collaborators: Carlos Pacheco, M.D., Patricia Louzon, PharmD., Robert Cambridge, D.O., Marcus Darrabie, M.D., Cheick Elmaali, M.D., Okorie Okorie, M.D. Jason Price, R.N., Sanjay Pattani, M.D., Brett Spenst, M.B.A., Amanda Tarkowski, M.D., Fahd Ali, M.D., Otsanya Ochogbu, PharmD., Bassel Raad, M.D., Mohammad Hmadeh, M.D., Mehul Patel, M.D.

## SUPPLEMENTAL INFORMATION

## Notes

### Competing Interest Statement

The authors have declared no competing interest.

### Funding Statement

Funding. This research received no specific grant from any funding agency in the public, commercial, or not-for-profit sectors. Expenses related to this publication are divided equally between the authors who are members of the division of critical care, infectious diseases and research institute at AdventHealth.

### Author Declarations

IRB Approval and Consent to participate: This protocol and end points for this observational retrospective study was evaluated and approved by the Institutional review board (IRB) of AdventHealth Central Florida Division. The research proposal, population involved and objectives of the study were also evaluated and approved by the Ethical committee of AdventHealth. Upon admission to hospital and/or ICU, patients or their family members agreed and signed appropriate consent to authorize physicians of the division of critical care as well as other consulting subspecialists to enroll them in different randomized trial for experimental therapies as wells as in observational/retrospective studies. Consent for publication: Patients and family members have agreed and signed appropriate consent to authorize AdventHealth Research Institute to access and utilize unidentified demographic and clinical information data saved in the electronic medical record and related only to the hospital admission due to COVID-19 for the purpose of clinical research, medical publications and/or medical education.

